# Wearable-derived cardiovascular fitness age and its lifestyle correlates in 442 adults

**DOI:** 10.64898/2026.03.20.26348891

**Authors:** Aditi Shanmugam, Kanika Gupta, Nihav Dhawale, Vatsal Singhal, Mohit Kumar, Bhuvan Srinivasan, Vinayak Narasimhan

## Abstract

Cardiovascular age is a powerful risk-communication tool that translates complex physiological data into an intuitive number, yet traditional estimates require clinical testing. Consumer wearables now estimate cardiorespiratory fitness age from photoplethysmography-derived heart rate data, enabling continuous, passive health monitoring, but whether such estimates capture substantive lifestyle variation has not been examined. We characterized Cardio Age, a wearable-derived cardiorespiratory fitness age estimate, in 442 Ultrahuman Ring users across a 12-month window ending February 2026, separating independent lifestyle correlates from direct or indirect algorithmic inputs. The mean Cardio Age gap (CA gap; mean Cardio Age minus chronological age) was −1.84 ± 2.97 years, with 82.6% of participants exhibiting younger estimated cardiovascular ages. Independent lifestyle metrics with no algorithmic link to Cardio Age showed significant associations: sleep efficiency (*r* = −0.194, *p <* 0.001), rapid eye movement (REM) sleep (*r* = −0.203, *p <* 0.001), sleep duration (*r* = −0.200, *p <* 0.001), and daily steps (*r* = −0.145, *p* = 0.003). A monotonic body mass index (BMI) dose-response was observed, with underweight participants showing a mean CA gap of −3.73 years versus −0.52 for obese participants. Extreme-group comparisons revealed that users with the youngest cardiovascular ages slept 37 minutes longer, achieved 22 more minutes of REM sleep, and had 1.8% higher sleep efficiency than those with the oldest cardiovascular ages (all *p <* 0.05). Sustained improvers over 12 months showed a mean CA reduction of 3.24 years, accompanied by decreased resting heart rate (− 0.8 bpm, *p <* 0.001) and increased estimated VO_2_ max (+1.3 mL/kg/min, *p <* 0.001), indicating that Cardio Age tracks physiological changes over time.

## Introduction

Cardiovascular age translates abstract risk-factor profiles into an intuitive metric that patients and the public can readily understand. The concept builds on the Framingham general cardiovascular risk profile^1^, which provided the foundation for subsequent vascular age frameworks that map risk factors onto an equivalent biological age and may improve risk communication relative to percentage-based scores^2,3^. Heart age has since been adopted in clinical guidelines and public health campaigns as a tool to personalize cardiovascular risk messaging^4,5^. A complementary approach, fitness age, derives biological age from cardiorespiratory fitness (CRF), typically measured or estimated as maximal oxygen uptake (VO_2_ max). Nes et al. developed a non-exercise prediction model for cardiorespiratory fitness in the HUNT Study^6^, and subsequently demonstrated that estimated CRF predicts all-cause and cardiovascular mortality in over 37,000 adults^7^. Large population studies have confirmed that CRF predicts mortality independently of traditional risk factors^8,9^, reinforcing CRF as a clinical vital sign^10^.

Traditionally, cardiovascular and fitness age estimation requires clinical exercise testing or laboratory-grade VO_2_ max measurement, limiting scalability. Consumer wearable devices primarily estimate VO_2_ max passively from photoplethysmography (PPG)-derived heart rate signals^11,12^, enabling daily monitoring without clinic visits. This has expanded the scope of wearable health monitoring beyond step counting to continuous physiological assessment^13^. Several commercial wearable platforms now translate wearable-derived estimated VO_2_ max into a fitness age metric. This shift from episodic clinical assessment to continuous passive monitoring enables users to track their cardiovascular fitness age daily, monitoring fundamental health metrics (resting heart rate, heart rate variability, and estimated VO_2_ max) through a single age-equivalent rather than abstract physiological values.

Prior validation studies have focused primarily on the accuracy of wearable heart rate and physiological measurements^14,15^, including wearable VO_2_ max estimates, where notable accuracy limitations have been reported^16^. Substantial evidence links lifestyle factors such as sleep duration^17,18^, body mass index (BMI)^19^, and physical activity^20^ to cardiovascular health. Although these factors are established determinants of cardiovascular health, whether they are captured by a wearable-derived fitness age metric, and whether such associations persist across observation windows, has not been systematically examined. To our knowledge, no study has tested whether within-person longitudinal trajectories of wearable cardiovascular age track measurable physiological changes. These gaps matter because wearable fitness age metrics are increasingly available to millions of consumers, and demonstrating that they reflect genuine physiological and lifestyle variation, not just algorithmic artifacts, is essential for establishing their scientific credibility and potential utility. A further analytical challenge is algorithmic circularity: because wearable fitness age is derived from VO_2_ max, which itself depends on resting heart rate, correlations between these constituent inputs and the output metric are partly tautological. Any characterization study must explicitly separate constituent from independent metrics to avoid overstating the novelty of circular associations.

Here we examined Cardio Age, a cardiorespiratory fitness age estimate derived from the Ultrahuman Ring, in 442 adult users across a 12-month observation window. Our aims were threefold: (a) characterize the distribution of the Cardio Age gap, defined as estimated cardiovascular age minus chronological age; (b) identify independent lifestyle correlates (metrics with no direct algorithmic link) to establish that Cardio Age captures physiological variation beyond its constituent inputs; and (c) examine within-person longitudinal trajectories to determine whether Cardio Age tracks real physiological improvements over time, supporting its utility as a practical tool for cardiovascular fitness monitoring.

## Results

### Study population and Cardio Age gap distribution

The study cohort comprised 442 Ultrahuman Ring users with a median age of 31 years (interquartile range [IQR] 26–42; range 18–77). The sample was 65.4% female (*N* = 289), 32.6% male (*N* = 144), and 2.0% other (*N* = 9). Median BMI was 25.0 kg/m^2^ (IQR 22.2–28.9), with 17 participants classified as underweight (3.8%), 207 as normal weight (46.8%), 126 as overweight (28.5%), and 92 as obese (20.8%). Self-reported mobility level was sedentary in 112 participants (25.3%), moderate in 272 (61.5%), and active in 58 (13.1%). The median number of observation days per user was 352 (IQR 267–361) for the 12-month window. All 442 users meeting the inclusion criterion (≥ 1 day of Cardio Age data) were included without further exclusion. Extreme groups were defined using the 10th and 90th percentiles of the 12-month CA gap distribution: the Youngest Hearts group (CA gap ≤− 5.4 years, *N* = 45) and the Oldest Hearts group (CA gap ≥ +2.3 years, *N* = 45). Full demographic characteristics are presented in Table 1.

**Table 1.** Demographic and clinical characteristics of the study cohort and extreme Cardio Age gap groups (12-month window). Youngest Hearts = CA gap ≤ −5.4 years (10th percentile); Oldest Hearts = CA gap ≥ +2.3 years (90th percentile). Values are median (IQR) for continuous variables and *N* (%) for categorical variables. *p*-values from Mann-Whitney *U* (continuous) or chi-square (categorical) tests comparing Youngest Hearts and Oldest Hearts groups.

| Variable | Full Cohort<br>($N = 442$) | Youngest Hearts<br>($N = 45$) | Oldest Hearts<br>($N = 45$) | $p$ |
| --- | --- | --- | --- | --- |
| Age, years | 31.0 (26.0, 42.0) | 29.0 (26.0, 33.0) | 35.0 (31.0, 45.0) | <0.001 |
| Gender |  |  |  | <0.001 |
| Female | 289 (65.4%) | 36 (80.0%) | 6 (13.3%) |  |
| Male | 144 (32.6%) | 4 (8.9%) | 39 (86.7%) |  |
| Other | 9 (2.0%) | 5 (11.1%) | 0 (0.0%) |  |
| BMI<sup>†</sup>, kg/m<sup>2</sup> | 25.0 (22.2, 28.9) | 22.8 (21.0, 26.2) | 29.6 (26.1, 33.3) | <0.001 |
| Weight<sup>†</sup>, kg | 73.5 (63.5, 86.0) | 69.0 (61.7, 77.7) | 89.0 (83.6, 100.0) | <0.001 |
| Height, cm | 170.0 (165.0, 177.0) | 173.0 (167.0, 175.0) | 175.0 (170.0, 183.0) | 0.178 |
| Mobility level |  |  |  | <0.001 |
| Sedentary | 112 (25.3%) | 3 (6.7%) | 17 (37.8%) |  |
| Moderate | 272 (61.5%) | 32 (71.1%) | 27 (60.0%) |  |
| Active | 58 (13.1%) | 10 (22.2%) | 1 (2.2%) |  |
| BMI category |  |  |  | <0.001 |
| Underweight (<18.5) | 17 (3.8%) | 4 (8.9%) | 0 (0.0%) |  |
| Normal (18.5–24.9) | 207 (46.8%) | 27 (60.0%) | 5 (11.1%) |  |
| Overweight (25–29.9) | 126 (28.5%) | 9 (20.0%) | 18 (40.0%) |  |
| Obese ($\geq 30$) | 92 (20.8%) | 5 (11.1%) | 22 (48.9%) | |
| CA gap, years | −2.01 (−3.46, −0.82) | −6.6 (−7.0, −5.9) | 4.5 (3.1, 5.8) | <0.001 |
| Observation days | 352.0 (267.0, 361.0) | 355.0 (333.0, 362.0) | 351.0 (227.0, 360.0) | 0.455 |

<sup>†</sup> BMI and weight are indirect constituents: known physiological determinants of VO<sub>2</sub> max (the algorithm's primary input).

The mean CA gap was −1.84 years (SD 2.97), with a median of −2.01 (IQR −3.46 to −0.82). A total of 82.6% of participants had negative CA gaps (younger estimated cardiovascular age), while 17.4% had positive gaps (older estimated cardiovascular age). The distribution was left-skewed (Figure 1). The preponderance of younger hearts likely reflects self-selection bias, as users who purchase and consistently wear a fitness-oriented ring are plausibly more health-conscious than the general population^21^.

**Figure 1.**
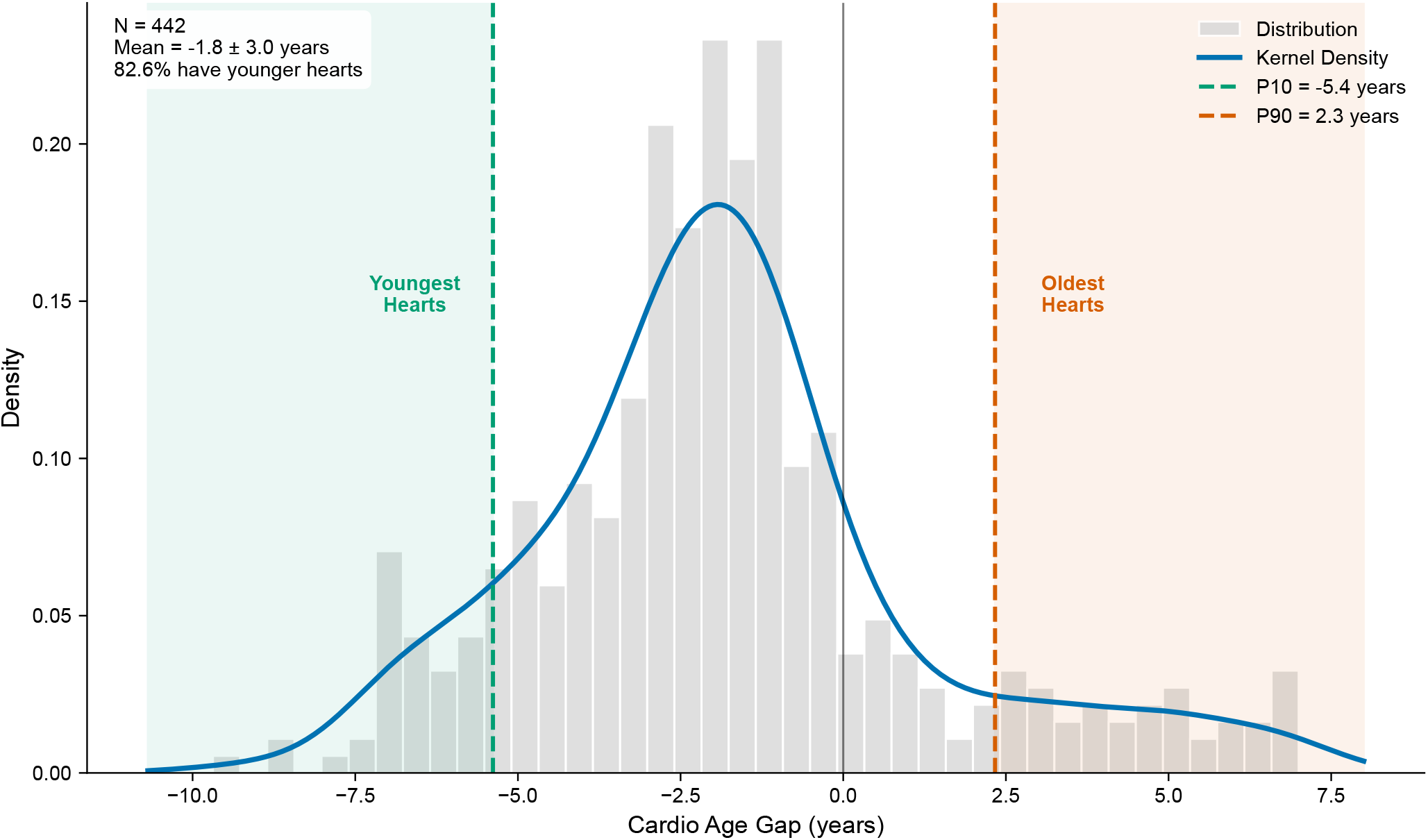
Distribution of the Cardio Age gap (mean Cardio Age minus chronological age, averaged over all available days within the 12-month window) for *N* = 442 participants. Histogram with kernel density estimate overlay. Vertical dashed lines indicate the 10th percentile (−5.4 years) and 90th percentile (+2.3 years) thresholds used to define the Youngest Hearts (*N* = 45) and Oldest Hearts (*N* = 45) extreme groups. Negative values indicate a cardiovascular system estimated to be younger than the individual’s chronological age. The distribution is left-skewed, with a mean of −1.84 ± 2.97 years and 82.6% of participants having negative gaps.

### Independent lifestyle correlates

To identify modifiable behaviors associated with cardiovascular fitness age beyond the algorithm’s direct inputs, we examined metrics with no direct algorithmic pathway. The Youngest Hearts group was 80.0% female, while the Oldest Hearts group was 86.7% male, consistent with sex-specific differences in the algorithm’s normative CRF-age equations^22^.

Among independent lifestyle metrics, the Youngest Hearts and Oldest Hearts groups differed significantly on sleep efficiency (90.4% vs. 88.6%, median difference +1.83, *p* = 0.002), REM sleep (120.5 vs. 98.3 min, +22.18, *p* = 0.007), sleep duration (454.6 vs. 417.4 min, +37.27, *p* = 0.032), and daily steps (7221 vs. 5540, +1681, *p* = 0.047). Deep sleep duration was not statistically significant (Supplementary Table S1; Figure 2).

**Figure 2.**
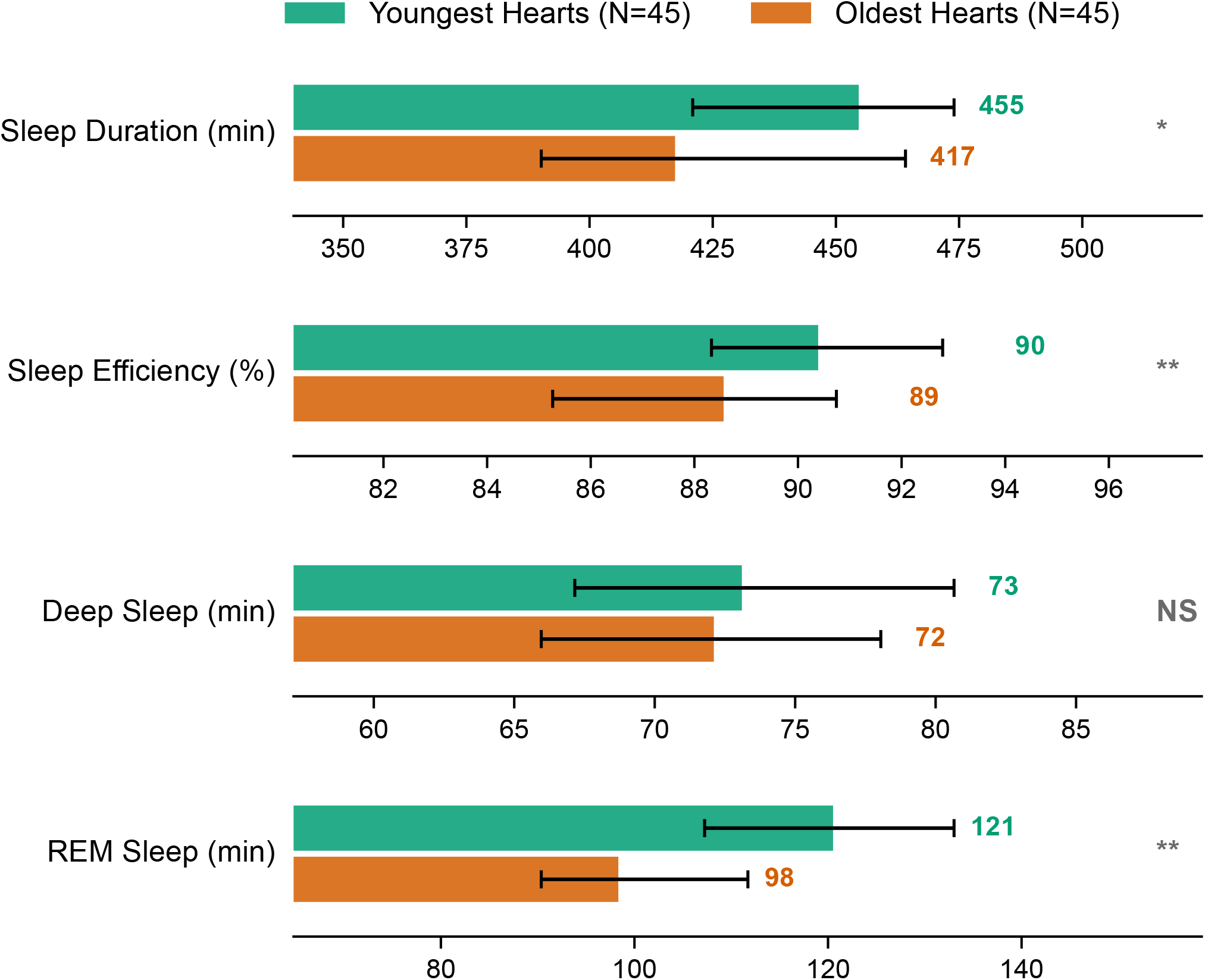
Sleep metrics in extreme Cardio Age gap groups (12-month window). Grouped bars comparing the Youngest Hearts (10th percentile, *N* = 45) and Oldest Hearts (90th percentile, *N* = 45) on independent sleep metrics. All comparisons by Mann-Whitney *U* tests. Sample sizes vary by metric due to data availability (sleep metrics: *N* = 42 vs. 39). ^**^ *p <* 0.01; * *p <* 0.05; NS, not significant.

Spearman rank correlations across the full cohort confirmed the extreme-group findings (Supplementary Table S2; Figure 3). The strongest independent correlates were REM sleep (*r* = −0.203, 95% CI −0.290 to −0.104, *p <* 0.001), sleep duration (*r* = −0.200, −0.287 to −0.097, *p <* 0.001), sleep efficiency (*r* = −0.194, −0.285 to −0.094, *p <* 0.001), and daily steps (*r* = −0.145, −0.240 to −0.048, *p* = 0.003). All three sleep metrics survived Bonferroni correction for 14 tests (*α/*14 = 0.004); daily steps also survived (*p* = 0.003). Deep sleep (*r* = −0.087, *p* = 0.088) was not significant. BMI (*r* = 0.278, 0.184–0.365, *p <* 0.001) and weight (*r* = 0.310, 0.227–0.393, *p <* 0.001) were strongly correlated but are classified as indirect constituents because BMI is a known physiological determinant of VO_2_ max (the CA algorithm’s primary input); these associations are therefore partly mediated through the algorithm. Recovery score (*r* = −0.320, −0.406 to −0.224, *p <* 0.001) was also strongly correlated but shares heart rate variability (HRV) and resting heart rate components with the algorithm.

**Figure 3.**
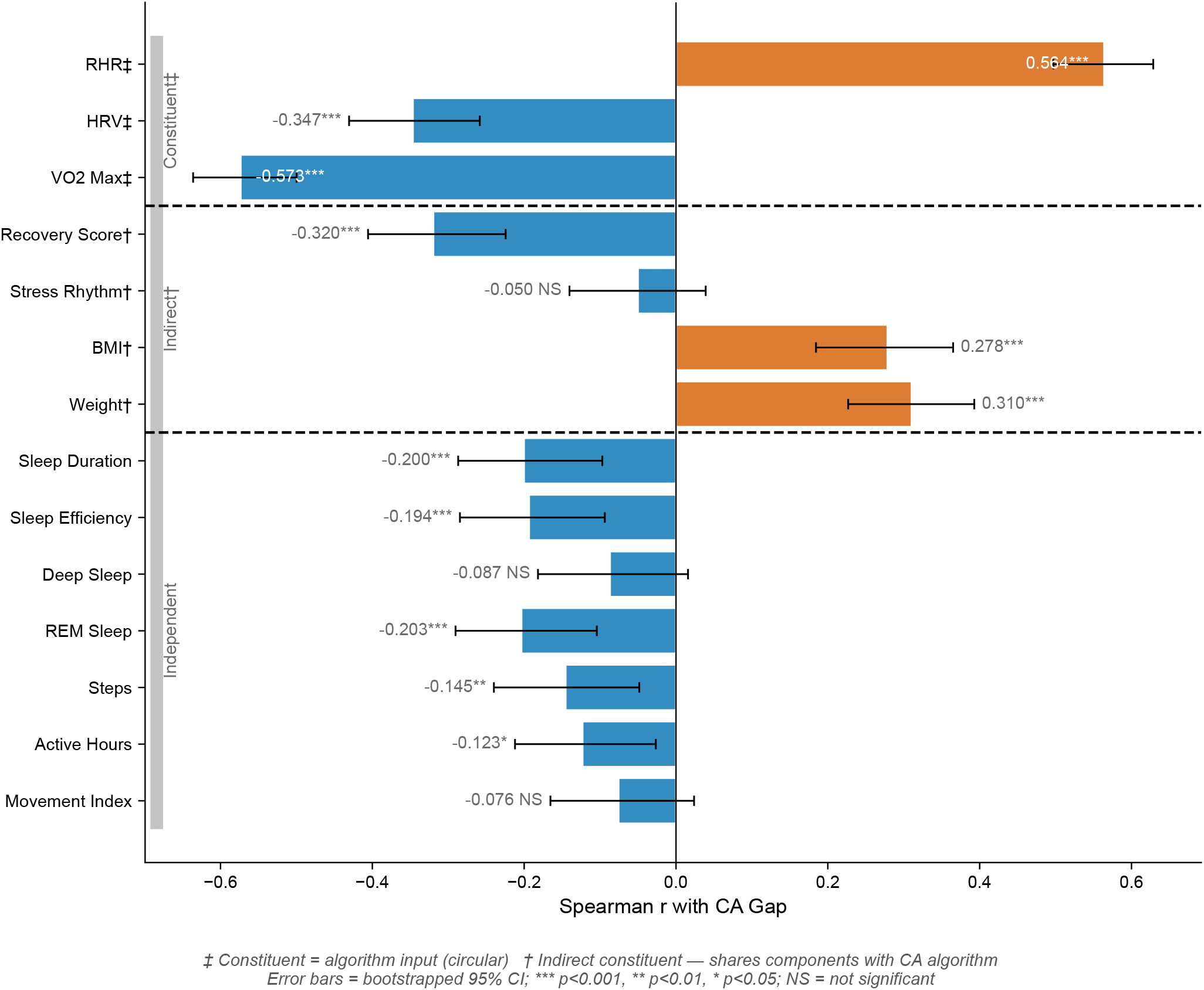
Spearman rank correlations between Cardio Age gap and wearable-derived metrics (12-month window). Horizontal bars show correlation coefficients. Metrics are grouped into three tiers: constituent (direct algorithm inputs, ^‡^), indirect constituent (shares algorithmic components, ^†^), and independent lifestyle correlates. Error bars represent bootstrapped 95% confidence intervals (1,000 iterations). ^***^ *p <* 0.001, ^**^ *p <* 0.01, ^*^ *p <* 0.05. NS, not significant.

A monotonic dose-response relationship was observed between BMI category and CA gap (Figure 4). Mean CA gap was −3.73 years for underweight participants (*N* = 17), −2.57 for normal weight (*N* = 207), −1.36 for overweight (*N* = 126), and −0.52 for obese (*N* = 92). Self-reported mobility level was also associated with CA gap: sedentary −1.01 years, moderate −1.84, active −3.48.

**Figure 4.**
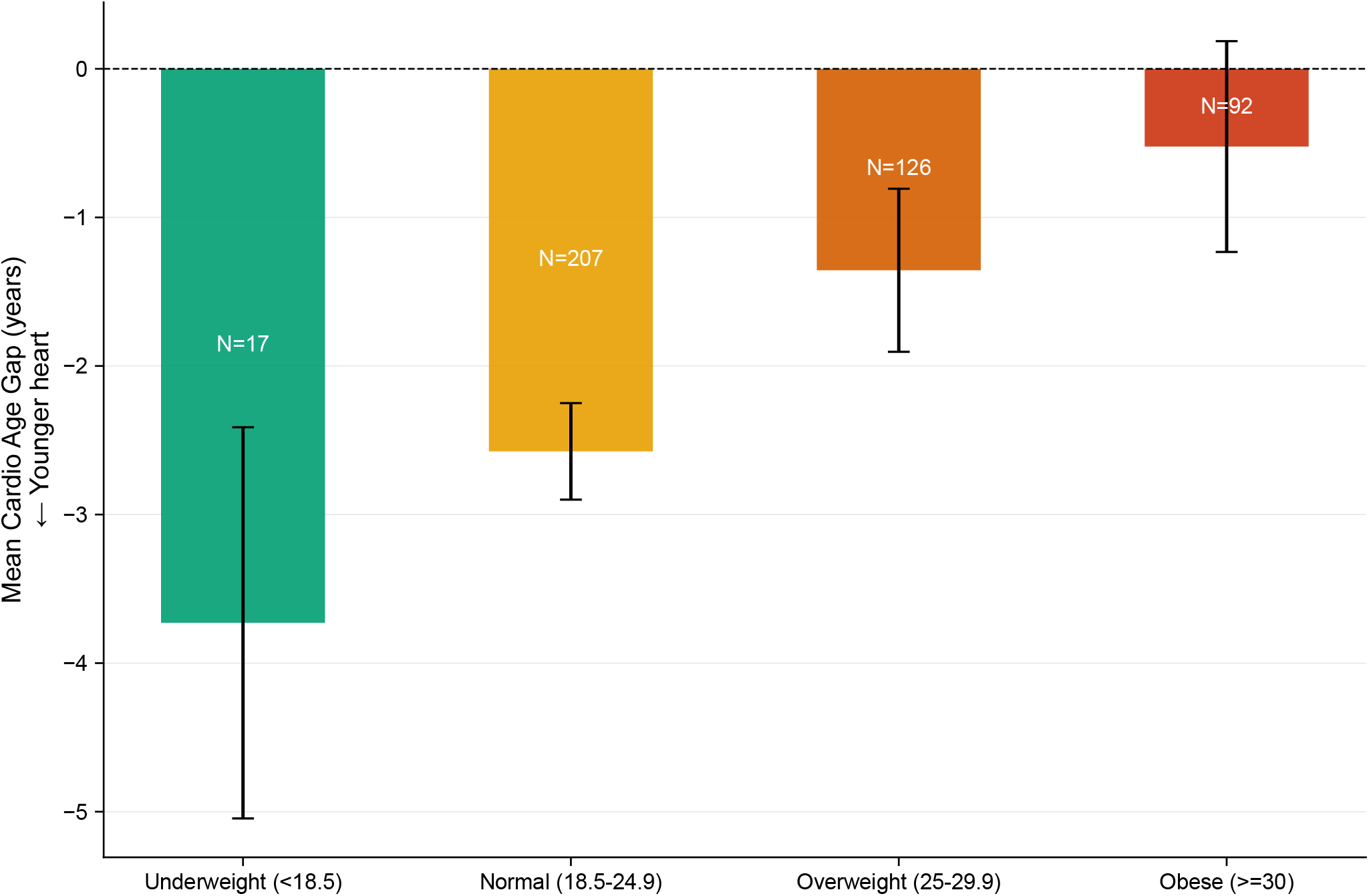
Body mass index dose-response relationship with Cardio Age gap (12-month window). Mean CA gap (±95% CI) by BMI^†^ category: underweight (*<*18.5 kg/m^2^, *N* = 17), normal weight (18.5–24.9, *N* = 207), overweight (25.0–29.9, *N* = 126), and obese (≥ 30.0, *N* = 92). ^†^BMI is not a direct algorithm input but is a known physiological determinant of VO_2_ max (the algorithm’s primary input); this association is therefore partly mediated through the algorithm.

### Constituent metric profiles and algorithmic transparency

The Cardio Age algorithm uses three tiers of inputs. Direct constituent inputs (flagged ^‡^) are the foundational algorithm inputs: nighttime resting heart rate, HRV root mean square of successive differences (RMSSD), and estimated VO_2_ max. Indirect constituents (flagged ^†^) are partly mediated through the algorithm: recovery score, stress rhythm score, BMI, and weight. Independent lifestyle correlates have no direct algorithmic link: sleep duration, sleep efficiency, deep sleep, REM sleep, daily steps, active hours, and movement index.

In extreme-group comparisons, the Youngest Hearts group had substantially lower nighttime resting heart rate^‡^ (47.9 vs. 61.7 bpm, *p <* 0.001), higher HRV^‡^ (RMSSD 55.9 vs. 37.2 ms, *p <* 0.001), and higher VO_2_ max^‡^ (48.7 vs. 36.4 mL/kg/min, *p <* 0.001). These differences are expected given the algorithmic relationship and are presented for completeness rather than as novel findings; they illustrate how Cardio Age translates abstract physiological metrics into a single age-equivalent. Recovery score^†^ also differed significantly (74.2 vs. 68.1, *p <* 0.001), as expected given its shared HRV and resting heart rate components. BMI^†^ (22.8 vs. 29.6 kg/m^2^, *p <* 0.001) and weight^†^ (69.0 vs. 89.0 kg, *p <* 0.001) also differed substantially between groups (Supplementary Table S1).

### Longitudinal trajectories

To examine whether within-person changes in Cardio Age correspond to physiological shifts, we classified users by their trajectory direction over 12 months using 30-day windows at the start and end of the observation period.

Using a threshold of |delta| *>* 2.0 years, 52 participants were classified as sustained large improvers (mean delta −3.24 years) and 59 as sustained large worseners (mean delta +3.49 years; Supplementary Tables S3–S4; Figure 5). The two groups were similar in age (33.1 vs. 35.4 years) and BMI (26.4 vs. 26.8 kg/m^2^). Improvements in Cardio Age were accompanied by decreased resting heart rate^‡^ (− 0.8 bpm vs. +1.1 bpm, *p <* 0.001) and increased VO_2_ max^‡^ (+1.3 vs. 0.8 mL/kg/min, *p <* 0.001). Temporal divergence in constituent metrics between improver and worsener groups became visible by months 3–4 (Supplementary Figure S1). Within-person Cardio Age was highly consistent over 12 months (median SD 1.21 years; 37.5% of users with SD *<* 1.0 years). The sustained improver cohort indicates that within-person Cardio Age changes correspond to measurable physiological shifts in resting heart rate and estimated VO_2_ max.

**Figure 5.**
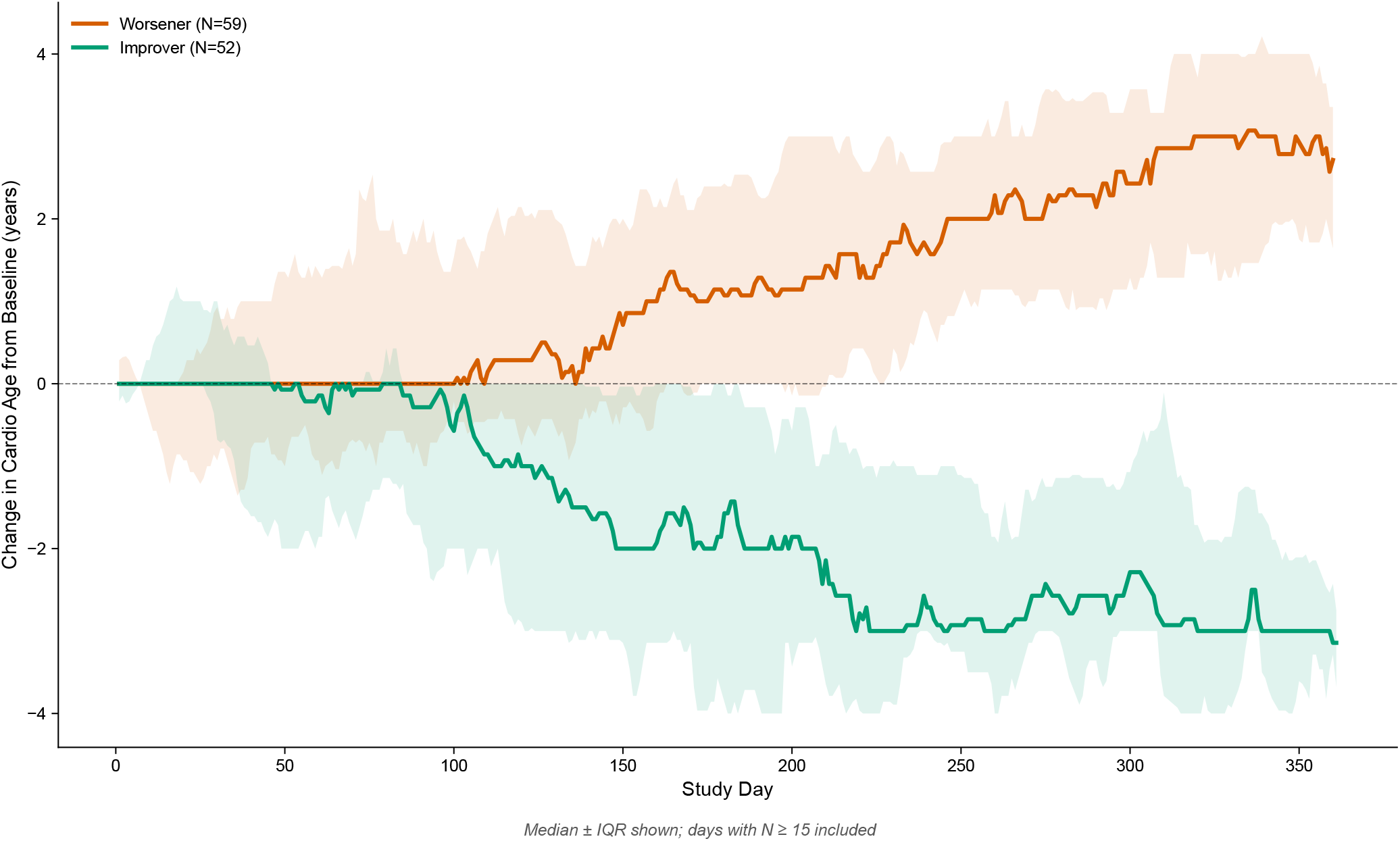
Twelve-month Cardio Age trajectories. Change from baseline (first 30-day mean) for sustained large improvers (delta *<* −2.0 years, *N* = 52) and sustained large worseners (delta *>* +2.0 years, *N* = 59). Lines represent group medians; shaded bands represent interquartile ranges. Sustained improvers showed a mean CA reduction of 3.24 years; sustained worseners showed a mean increase of 3.49 years over the 12-month period.

## Discussion

We characterized the distribution, lifestyle correlates, and longitudinal trajectories of Cardio Age, a wearable-derived cardiorespiratory fitness age estimate, in 442 Ultrahuman Ring users across a 12-month observation window. The principal findings were that the strongest independent correlates of the Cardio Age gap were REM sleep, sleep duration, sleep efficiency, and daily steps, all metrics with no direct algorithmic link to Cardio Age. BMI and weight, classified as indirect constituents, showed associations consistent with prior epidemiological findings. Within-person Cardio Age was highly consistent over time (median SD 1.21 years). Sustained improvers demonstrated meaningful reductions in resting heart rate and increases in estimated VO_2_ max, indicating that Cardio Age reflects physiological changes over time.

Although numerous cardiovascular age concepts exist, several features support the relevance of the Ultrahuman Cardio Age metric specifically. Cardiorespiratory fitness, as quantified by VO_2_ max, is a well-documented independent predictor of all-cause and cardiovascular mortality^7–9^. An algorithm that estimates VO_2_ max from wearable data therefore offers a practical bridge between clinical fitness testing and everyday wellness monitoring. The Ultrahuman CA algorithm draws on established cardiorespiratory fitness estimation frameworks^22,23^, adapting laboratory-grade physiological foundations for a consumer wearable context. The cross-sectional correlates we observe (sleep, BMI as indirect constituent) align with known determinants of cardiorespiratory fitness in the epidemiological literature, providing face validity. Within-person stability (median SD 1.21 years over 12 months) confirms the metric captures genuine individual-level physiological variation rather than measurement noise.

The observed associations are consistent with known physiological relationships. The inverse relationship between BMI and CRF is well documented^8,20,24^, higher BMI is associated with increased lifetime cardiovascular risk^19^, and our finding that higher BMI is associated with older Cardio Age extends these relationships to a consumer wearable context. Sleep duration has been linked to cardiovascular health in large prospective studies; short sleep increases the risk of all-cause mortality and coronary heart disease^17,18,25^. The association between REM sleep and younger Cardio Age is consistent with evidence linking sleep architecture to autonomic regulation and cardiovascular health^26^. Recovery score, a composite autonomic measure incorporating resting heart rate and HRV, was classified as an indirect constituent because it shares algorithmic components with Cardio Age. Its strong association with CA gap (*r* = −0.320) aligns with a growing literature on HRV-based recovery monitoring^27,28^.

The fitness age concept underlying our work was established by Nes et al., who demonstrated that estimated cardiorespiratory fitness predicts all-cause mortality in over 37,000 adults^6,7^. Our findings extend this framework from single-timepoint clinical assessments to continuous daily monitoring, providing evidence that a wearable-derived fitness age metric reflects lifestyle and physiological variation beyond its algorithmic inputs over a 12-month period.

Algorithmic circularity is an important consideration for interpreting these results. Nighttime resting heart rate is a key input to the Cardio Age algorithm, which estimates VO_2_ max using a proprietary combination of validated methods^22,23^. Correlations between these constituent metrics and CA gap are therefore expected, and we present them as descriptive characterization.

BMI is an indirect constituent: it is not a direct algorithm input, but BMI is a known physiological determinant of VO_2_ max^8^, so the BMI-CA gap association is partly mediated through the algorithm’s primary input. After flagging these layers, the remaining independent correlates (sleep duration, REM sleep, sleep efficiency, and daily steps) represent associations with no direct algorithmic link. The fact that these independent lifestyle metrics correlated with CA gap despite having no algorithmic pathway suggests that Cardio Age captures physiological variation beyond its constituent inputs. These associations were also stable across 3-, 6-, and 12-month observation windows, with sleep duration, sleep efficiency, and REM sleep showing consistent correlation magnitudes across all three horizons. A multivariate model including BMI, sleep efficiency, gender, and age explained 30.1% of CA gap variance (Supplementary Table S5).

A moderate positive correlation was observed between CA gap and chronological age (*r* = 0.238, *p <* 0.001), indicating that older participants tended to have more positive (less favorable) CA gaps. This pattern could reflect genuine age-related cardiovascular fitness decline, or it could indicate calibration differences in the normative CRF-age reference data^22^ relative to this cohort’s demographic composition. The two explanations are not mutually exclusive; disentangling self-selection from algorithmic calibration would require external validation against laboratory-measured VO_2_ max, which is beyond the scope of this characterization study.

Sustained trajectories over 12 months demonstrated that Cardio Age tracks consistent physiological trends: improvements were accompanied by decreased resting heart rate and increased estimated VO_2_ max, with divergence between improver and worsener groups becoming visible by months 3–4. The physiological mechanism most plausibly captured by Cardio Age is autonomic adaptation to aerobic exercise. Regular endurance training increases vagal tone, reducing resting heart rate^29^, a key input to the Cardio Age algorithm. This autonomic adaptation pathway is consistent with the trajectory patterns observed and is consistent with Cardio Age functioning as a continuous fitness metric that reflects cardiovascular conditioning. Sleep duration may correlate cross-sectionally with CA gap as a marker of overall health status (individuals who sleep more may have lower allostatic load and more time for exercise) rather than as a direct driver of resting heart rate changes.

The sustained worsener group (*N* = 59, mean delta +3.49 years) was slightly larger than the improver group (*N* = 52) and was 61% female compared with 50% in improvers. Several mechanisms could underlie worsening trajectories: genuine physiological decline (e.g., reduced physical activity, weight gain, or increased allostatic load), regression to the mean for participants who started with unusually favorable CA gap values, or seasonal or behavioral changes over time. The two groups were demographically similar at baseline (age 33.1 vs. 35.4 years; BMI 26.4 vs. 26.8 kg/m^2^), suggesting that worsening was not simply a function of older or less fit starting points. This symmetric treatment of worsening and improving trajectories strengthens the interpretation that Cardio Age tracks bidirectional physiological change.

Heart age has been proposed as a motivational tool for health behavior change^2,5^, but evidence has been limited to single-timepoint clinical assessments. Cardio Age extends this concept by providing continuous, daily feedback: the sustained improver cohort achieved a mean 3.24-year reduction in cardiovascular fitness age over 12 months without any structured intervention, suggesting that passive daily feedback on fitness age may itself sustain user engagement with cardiovascular health. Cardio Age also addresses a gap in how individuals interact with their health data. A decrease in resting heart rate of 0.8 bpm or an increase in estimated VO_2_ max of 1.3 mL/kg/min is difficult for the average user to interpret or act upon; by contrast, a three-year decrease in cardiovascular age provides an immediately intuitive signal. The translation of abstract physiological values into a personal, age-based metric is what may make Cardio Age actionable at scale. When users can see that their daily behaviors (sleep habits, physical activity, weight management) are reflected in a single number they can track over time, the perceived feedback loop between behavior and outcome becomes tangible, potentially supporting sustained engagement with personalized health monitoring. The dose-response relationship between BMI and CA gap, the significant associations with sleep metrics and daily steps, and the trajectory data showing sustained improvement all point to a metric that may serve as a practical indicator for everyday cardiovascular health awareness at the population level.

Several methodological strengths support confidence in these findings: daily longitudinal data from 442 users rather than single-timepoint assessment, transparent separation of constituent and independent metrics, and within-person stability analysis over 12 months. Limitations include: (1) *Algorithmic circularity*: constituent metrics are direct or indirect algorithm inputs, as fully disclosed above. (2) *Sample characteristics*: participants are self-selected wearable users (median age 31 years) with self-reported anthropometrics; however, these characteristics are representative of the early-adopter population to whom the metric is delivered. (3) *Observational design*: associations do not imply causation; however, within-person longitudinal analyses and concordance with prior epidemiological evidence provide convergent support. (4) *External validation*: no comparison to laboratory-measured VO_2_ max or CPET-derived fitness age was performed; this work should be interpreted as a characterization study establishing that the metric reflects genuine physiological and lifestyle variation.

Future work should pursue prospective validation linking Cardio Age to cardiovascular outcomes, algorithm refinement incorporating sleep and recovery directly, cross-device generalizability studies, and examination of intervention effects on Cardio Age trajectories.

## Methods

### Study design and participants

This was a retrospective observational cohort study of 442 adult users of the Ultrahuman Ring AIR smart ring. Inclusion criteria were age ≥18 years and at least one day of Cardio Age data between March 1, 2025, and February 28, 2026. One participant aged *<*18 was excluded from the initial dataset of 443 users. The primary analysis window was 12 months (March 1, 2025, to February 28, 2026; 365 days). Demographic data, including age, sex, height, weight, and self-reported mobility level, were collected while onboarding on the Ultrahuman platform.

### Ethics statement

This was a real-world, retrospective, observational study based on data derived from Ultrahuman platform users and adhered to Ultrahuman’s terms of use^30^ and privacy policy^31^, which allows for analysis of de-identified grouped data for scientific research. Participants consented via the onboarding process on the Ultrahuman platform and continued product use. As the study was non-invasive and involved no dietary, sleep, or exercise interventions, with all reported data de-identified, explicit institutional ethics board approval was not required. A separate set of analysts extracted the data, ran computational approaches, and then reviewed the results to ensure blinding.

### Device

The Ultrahuman Ring AIR is a titanium finger-worn smart ring equipped with a green-light photoplethysmography (PPG) sensor for continuous heart rate monitoring, a tri-axis accelerometer for motion and activity detection, and a skin temperature sensor. The ring form factor enables continuous, unobtrusive physiological data collection during daily activities and sleep without requiring user interaction.

### Cardio Age algorithm

Cardio Age is a proprietary cardiorespiratory fitness age estimate. Estimated VO_2_ max is derived from a proprietary weighted combination of established cardiorespiratory fitness estimation methods^22,23^, incorporating nighttime resting heart rate as a foundational input. The estimated VO_2_ max is then converted to a fitness age using sex-specific normative CRF-age data^22^, with temporal smoothing applied to limit day-to-day variation. Cardio Age is estimated daily for each user. Self-reported mobility level influences the estimation as an adjustment factor. These relationships create layers of algorithmic circularity that are explicitly addressed in our analytical framework.

### Outcome and predictor variables

The primary outcome was the Cardio Age gap (CA gap), defined as the mean of all available daily Cardio Age values across the observation window minus chronological age. Negative values indicate a cardiovascular system estimated to be younger than chronological age. Per-user values for each wearable-derived metric were computed as the mean of all available daily observations within the observation window.

Constituent metrics (direct algorithm inputs, flagged with ^‡^ throughout) were nighttime resting heart rate (bpm), HRV RMSSD (ms), and estimated VO_2_ max (mL/kg/min). Indirect constituent metrics (flagged with ^†^) were recovery score, stress rhythm score, BMI (kg/m^2^), and weight (kg). Recovery score and stress rhythm score share HRV and resting heart rate components with the Cardio Age algorithm. BMI and weight are not direct algorithm inputs but are known physiological determinants of VO_2_ max^8^, so associations between these metrics and CA gap are partly mediated through the algorithm’s primary input.

Independent lifestyle correlates included sleep duration (min), sleep efficiency (%), deep sleep duration (min), REM sleep duration (min), daily steps, active hours, and movement index.

Sleep efficiency was defined as the ratio of total sleep time to total time in bed, expressed as a percentage. Ultrahuman-proprietary composite metrics were defined as follows. Recovery score (0–100) is a platform-derived composite incorporating HRV, resting heart rate, sleep quality, and prior-day activity; because it shares HRV and resting heart rate components with the Cardio Age algorithm, it was classified as an indirect constituent^†^. Stress rhythm score (0–100) reflects autonomic balance patterns and similarly shares algorithmic inputs with Cardio Age^†^. Movement index is a composite activity metric derived from accelerometer data reflecting overall daily movement patterns. These metrics are platform-derived composites and should not be equated with validated clinical instruments.

Extreme groups were defined using the 10th and 90th percentiles of the 12-month CA gap distribution: Youngest Hearts (CA gap ≤− 5.4 years, *N* = 45) and Oldest Hearts (CA gap ≥ +2.3 years, *N* = 45). This analytical choice enables comparison of users with the most divergent cardiovascular fitness ages.

Longitudinal change was quantified as delta CA: last 30-day mean minus first 30-day mean CA over the 12-month window. Thirty-day windows were used to provide robust estimates and mitigate regression to the mean. Improvers were defined as delta *<* −2.0 years; worseners as delta *>* +2.0. These thresholds were chosen to exceed the within-person measurement variability (median SD 1.21 years), ensuring that classified changes exceed typical metric noise.

### Statistical analysis

Continuous variables are reported as median (IQR); categorical variables as *N* (%). Extreme-group comparisons used Mann-Whitney *U* tests (two-tailed) with median difference and 95% confidence interval. Spearman rank correlations were computed with bootstrapped 95% confidence intervals (1,000 iterations). Longitudinal within-person changes were assessed by Wilcoxon signed-rank tests within trajectory groups and Kruskal-Wallis tests between trajectory groups. Recovery score sentinel values (−1) were converted to missing before aggregation. Statistical significance was set at *α* = 0.05 (two-tailed). For the 14 correlations reported in Supplementary Table S2 (constituent, indirect constituent, and independent metrics), a Bonferroni-corrected threshold of *α/*14 = 0.004 was applied; results are reported at both nominal and corrected thresholds. Exact *p*-values are reported throughout. Analyses were performed in Python 3.12 (pandas 2.2^32^, scipy 1.12^33^, statsmodels 0.14^34^, scikit-learn 1.4^35^).

### Use of artificial intelligence

Artificial intelligence tools (Claude, Anthropic) were used to assist with analysis scripting and copyediting of draft text. All code was verified by the authors; all statistical outputs and interpretations were independently reviewed and approved by all authors, who take full responsibility for the accuracy and integrity of the work.

## Data Availability

Summary statistics and analysis outputs supporting the findings of this study are provided in the manuscript and Supplementary Information. Individual-level data are proprietary to Ultrahuman Healthcare Pvt Ltd and are not publicly available due to user privacy restrictions.

## Acknowledgements

The authors thank the Ultrahuman Ring users whose de-identified data made this study possible.

## Author contributions statement

V.N. conceived and designed the study, supervised the analysis, and co-wrote the manuscript. A.S. co-designed the study, performed data extraction and curation, and co-wrote the manuscript. K.G. and N.D. contributed to interpretation of results and co-wrote the manuscript. V.S., M.K., and B.S. contributed to manuscript preparation and revision. All authors reviewed and approved the final manuscript.

## References

1. D’Agostino, R. B. et al. General cardiovascular risk profile for use in primary care: the Framingham Heart Study. Circulation 117, 743–753, 10.1161/CIRCULATIONAHA.107.699579 (2008).

2. Groenewegen, K. A. et al. Vascular age to determine cardiovascular disease risk: A systematic review of its concepts, definitions, and clinical applications. Eur. J. Prev. Cardiol. 23, 264–274, 10.1177/2047487314566999 (2016).

3. Cuende, J. I. Vascular age versus cardiovascular risk: Clarifying concepts. Rev. Esp. Cardiol. 69, 243–246, 10.1016/j.rec.2015.10.019 (2016).

4. Piepoli, M. F. et al. 2016 European guidelines on cardiovascular disease prevention in clinical practice. Eur. Heart J. 37, 2315–2381, 10.1093/eurheartj/ehw106 (2016).

5. Bonner, C. et al. Interventions using heart age for cardiovascular disease risk communication: systematic review of psychological, behavioral, and clinical effects. JMIR Cardio 5, e31056, 10.2196/31056 (2021).

6. Nes, B. M. et al. Estimating V O2peak from a nonexercise prediction model: the HUNT Study, Norway. Med. Sci. Sports Exerc. 43, 2024–2030, 10.1249/MSS.0b013e31821d3f6f (2011).

7. Nes, B. M., Vatten, L. J., Nauman, J., Janszky, I. & Wisløff, U. A simple nonexercise model of cardiorespiratory fitness predicts long-term mortality. Med. Sci. Sports Exerc. 46, 1159–1165, 10.1249/MSS.0000000000000219 (2014).

8. Ross, R. et al. Importance of assessing cardiorespiratory fitness in clinical practice: a case for fitness as a clinical vital sign. Circulation 134, e653–e699, 10.1161/CIR.0000000000000461 (2016).

9. Kodama, S. et al. Cardiorespiratory fitness as a quantitative predictor of all-cause mortality and cardiovascular events in healthy men and women: a meta-analysis. JAMA 301, 2024–2035, 10.1001/jama.2009.681 (2009).

10. Kaminsky, L. A. et al. The importance of cardiorespiratory fitness in the United States: the need for a national registry. Circulation 127, 652–662, 10.1161/CIR.0b013e31827ee100 (2013).

11. Allen, J. Photoplethysmography and its application in clinical physiological measurement. Physiol. Meas. 28, R1–R39, 10.1088/0967-3334/28/3/R01 (2007).

12. Shcherbina, A. et al. Accuracy in wrist-worn, sensor-based measurements of heart rate and energy expenditure in a diverse cohort. J. Pers. Med. 7, 3, 10.3390/jpm7020003 (2017).

13. Dunn, J., Runge, R. & Snyder, M. Wearables and the medical revolution. Per. Med. 15, 429–448, 10.2217/pme-2018-0044 (2018).

14. Bent, B., Goldstein, B. A., Kibbe, W. A. & Dunn, J. P. Investigating sources of inaccuracy in wearable optical heart rate sensors. NPJ Digit. Med. 3, 18, 10.1038/s41746-020-0226-6 (2020).

15. Fuller, D. et al. Reliability and validity of commercially available wearable devices for measuring steps, energy expenditure, and heart rate: systematic review. JMIR Mhealth Uhealth 8, e18694, 10.2196/18694 (2020).

16. Passler, S., Bohrer, J., Blochinger, L. & Senner, V. Validity of wrist-worn activity trackers for estimating VO2max and energy expenditure. Int. J. Environ. Res. Public Health 16, 3037, 10.3390/ijerph16173037 (2019).

17. Cappuccio, F. P., D’Elia, L., Strazzullo, P. & Miller, M. A. Sleep duration and all-cause mortality: a systematic review and meta-analysis of prospective studies. Sleep 33, 585–592, 10.1093/sleep/33.5.585 (2010).

18. St-Onge, M.-P. et al. Sleep duration and quality: impact on lifestyle behaviors and cardiometabolic health: a scientific statement from the American Heart Association. Circulation 134, e367–e386, 10.1161/CIR.0000000000000444 (2016).

19. Khan, S. S. et al. Association of body mass index with lifetime risk of cardiovascular disease and compression of morbidity. JAMA Cardiol. 3, 280–287, 10.1001/jamacardio.2018.0022 (2018).

20. Lavie, C. J., Ozemek, C., Carbone, S., Katzmarzyk, P. T. & Blair, S. N. Sedentary behavior, exercise, and cardiovascular health. Circ. Res. 124, 799–815, 10.1161/CIRCRESAHA.118.312669 (2019).

21. Piwek, L., Ellis, D. A., Andrews, S. & Joinson, A. The rise of consumer health wearables: promises and barriers. PLoS Med. 13, e1001953, 10.1371/journal.pmed.1001953 (2016).

22. Jackson, A. S., Sui, X., Hébert, J. R., Church, T. S. & Blair, S. N. Role of lifestyle and aging on the longitudinal change in cardiorespiratory fitness. Arch. Intern. Med. 169, 1781–1787, 10.1001/archinternmed.2009.312 (2009).

23. Tanaka, H., Monahan, K. D. & Seals, D. R. Age-predicted maximal heart rate revisited. J. Am. Coll. Cardiol. 37, 153–156, 10.1016/S0735-1097(00)01054-8 (2001).

24. Wei, M. et al. Relationship between low cardiorespiratory fitness and mortality in normal-weight, overweight, and obese men. JAMA 282, 1547–1553, 10.1001/jama.282.16.1547 (1999).

25. Lao, X. Q. et al. Sleep quality, sleep duration, and the risk of coronary heart disease: a prospective cohort study with 60,586 adults. J. Clin. Sleep Med. 14, 109–117, 10.5664/jcsm.6894 (2018).

26. Tobaldini, E. et al. Sleep, sleep deprivation, autonomic nervous system and cardiovascular diseases. Neurosci. Biobehav. Rev. 74, 321–329, 10.1016/j.neubiorev.2016.07.004 (2017).

27. Shaffer, F. & Ginsberg, J. P. An overview of heart rate variability metrics and norms. Front. Public Health 5, 258, 10.3389/fpubh.2017.00258 (2017).

28. Plews, D. J., Laursen, P. B., Stanley, J., Kilding, A. E. & Buchheit, M. Training adaptation and heart rate variability in elite endurance athletes: opening the door to effective monitoring. Sports Med. 43, 773–781, 10.1007/s40279-013-0071-8 (2013).

29. Carter, J. B., Banister, E. W. & Blaber, A. P. Effect of endurance exercise on autonomic control of heart rate. Sports Med. 33, 33–46, 10.2165/00007256-200333010-00003 (2003).

30. Ultrahuman. Terms of use. https://ultrahumanapp.notion.site/TERMS-OF-USE-7a541a34b77b4cb79566a327578873bb (2024). Accessed: 2026-03-20.

31. Ultrahuman. Privacy policy. https://ultrahumanapp.notion.site/PRIVACY-POLICY-16dccc2b1b2b4c4bb9a462c155b3bef1 (2024). Accessed: 2026-03-20.

32. McKinney, W. Data structures for statistical computing in Python. In Proceedings of the 9th Python in Science Conference, 56–61, 10.25080/Majora-92bf1922-00a (2010).

33. Virtanen, P. et al. SciPy 1.0: Fundamental algorithms for scientific computing in Python. Nat. Methods 17, 261–272, 10.1038/s41592-019-0686-2 (2020).

34. Seabold, S. & Perktold, J. statsmodels: Econometric and statistical modeling with Python. In Proceedings of the 9th Python in Science Conference, 92–96, 10.25080/Majora-92bf1922-011 (2010).

35. Pedregosa, F. et al. Scikit-learn: Machine learning in Python. J. Mach. Learn. Res. 12, 2825–2830 (2011).

