## Supplementary Information for "Wearable-derived cardiovascular fitness age and its lifestyle correlates in 442 adults"

**Table S1.** All wearable-derived metrics in extreme Cardio Age gap groups (12-month window). Values are median (IQR). $p$-values from Mann-Whitney $U$ tests. Metrics are grouped by type: constituent (‡, direct algorithm inputs), indirect constituent (†, shares algorithmic components), and independent (no algorithmic link).

| Metric | Type | Youngest Hearts | Oldest Hearts | Median Diff | $p$ |
|---|---|---|---|---|---|
| RHR (bpm)‡ | Constituent | 47.9 (45.4, 49.4) | 61.7 (59.2, 66.4) | −13.78 | <0.001 |
| HRV RMSSD (ms)‡ | Constituent | 55.9 (46.4, 68.9) | 37.2 (32.1, 47.6) | +18.63 | <0.001 |
| $VO_2$ max (mL/kg/min)‡ | Constituent | 48.7 (45.3, 51.6) | 36.4 (33.7, 38.6) | +12.32 | <0.001 |
| Recovery score† | Indirect constituent | 74.2 (69.9, 76.9) | 68.1 (63.1, 71.0) | +6.08 | <0.001 |
| Stress rhythm† | Indirect constituent | 77.9 (73.3, 80.6) | 77.5 (69.8, 81.8) | +0.38 | 0.759 |
| BMI (kg/m$^2$)† | Indirect constituent | 22.8 (21.0, 26.2) | 29.6 (26.1, 33.3) | −6.78 | <0.001 |
| Weight (kg)† | Indirect constituent | 69.0 (61.7, 77.7) | 89.0 (83.6, 100.0) | −20.00 | <0.001 |
| Sleep duration (min) | Independent | 454.6 (420.9, 474.0) | 417.4 (390.2, 464.1) | +37.27 | 0.032 |
| Sleep efficiency (%) | Independent | 90.4 (88.3, 92.8) | 88.6 (85.3, 90.7) | +1.83 | 0.002 |
| Deep sleep (min) | Independent | 73.1 (67.2, 80.7) | 72.1 (66.0, 78.0) | +1.00 | 0.427 |
| REM sleep (min) | Independent | 120.5 (107.2, 133.0) | 98.3 (90.4, 111.7) | +22.18 | 0.007 |
| Daily steps | Independent | 7221 (5171, 8816) | 5540 (4074, 7727) | +1681 | 0.047 |
| Active hours | Independent | 8.4 (6.2, 9.8) | 7.0 (5.9, 8.4) | +1.42 | 0.059 |
| Movement index | Independent | 70.8 (62.2, 76.5) | 69.2 (56.7, 77.3) | +1.60 | 0.340 |

‡ Direct constituent (algorithm input). †Indirect constituent; shares algorithmic components.
Youngest Hearts: CA gap $\leq -5.4$ years ($N = 45$); Oldest Hearts: CA gap $\geq +2.3$ years ($N = 45$). Sample sizes vary by metric due to data availability.

**Table S2.** Spearman rank correlations between Cardio Age gap and all wearable-derived metrics (12-month window). Bootstrapped 95% confidence intervals from 1,000 iterations.

| Metric | Type | $r$ | 95% CI | $N$ | $p$ |
|---|---|---|---|---|---|
| RHR‡ | Constituent | 0.564 | (0.497, 0.629) | 442 | <0.001 |
| HRV‡ | Constituent | −0.347 | (−0.431, −0.258) | 442 | <0.001 |
| $VO_2$ max‡ | Constituent | −0.573 | (−0.636, −0.500) | 428 | <0.001 |
| Recovery score† | Indirect constituent | −0.320 | (−0.406, −0.224) | 442 | <0.001 |
| Stress rhythm† | Indirect constituent | −0.050 | (−0.140, 0.039) | 442 | 0.293 |
| BMI† | Indirect constituent | 0.278 | (0.184, 0.365) | 442 | <0.001 |
| Weight† | Indirect constituent | 0.310 | (0.227, 0.393) | 442 | <0.001 |
| Sleep duration | Independent | −0.200 | (−0.287, −0.097) | 387 | <0.001 |
| Sleep efficiency | Independent | −0.194 | (−0.285, −0.094) | 387 | <0.001 |
| Deep sleep | Independent | −0.087 | (−0.182, 0.016) | 387 | 0.088 |
| REM sleep | Independent | −0.203 | (−0.290, −0.104) | 387 | <0.001 |
| Steps | Independent | −0.145 | (−0.240, −0.048) | 430 | 0.003 |
| Active hours | Independent | −0.123 | (−0.212, −0.027) | 430 | 0.011 |
| Movement index | Independent | −0.076 | (−0.165, 0.024) | 432 | 0.116 |

‡ Direct constituent (algorithm input). †Indirect constituent; partly mediated through the algorithm.

**Table S3.** Twelve-month sustained trajectory profiles. Sustained improvers (delta $< -2.0$ years) and sustained worseners (delta $> +2.0$ years).

| Characteristic | Sustained Improvers ($N = 52$) | Sustained Worseners ($N = 59$) |
|---|---|---|
| Mean age, years | 33.1 | 35.4 |
| Mean BMI, kg/m$^2$ | 26.4 | 26.8 |
| Female, % | 50.0 | 61.0 |
| Mean delta CA, years | $-3.24$ | $+3.49$ |

**Table S4.** Between-group longitudinal metric changes for 12-month sustained trajectory groups. $p$-values from Kruskal-Wallis tests.

| Metric | Type | Improver Δ | Worsener Δ | $p$ |
|---|---|---|---|---|
| RHR (bpm)‡ | Constituent | $-0.84$ | $+1.05$ | <0.001 |
| HRV RMSSD (ms)‡ | Constituent | $+1.05$ | $-1.89$ | <0.001 |
| $VO_2$ max (mL/kg/min)‡ | Constituent | $+1.26$ | $-0.80$ | <0.001 |
| Recovery score† | Indirect constituent | $+2.49$ | $+1.49$ | 0.157 |
| Sleep duration (min) | Independent | $+5.1$ | $-38.7$ | 0.465 |
| Sleep efficiency (%) | Independent | $+0.58$ | $-0.45$ | 0.273 |
| Deep sleep (min) | Independent | $-4.04$ | $-16.20$ | 0.465 |
| REM sleep (min) | Independent | $-3.07$ | $-22.91$ | 0.361 |

**Table S5.** Multivariate linear regression of CA gap on independent predictors (12-month window). Primary model includes BMI†, sleep duration, sleep efficiency, gender, and age. Sensitivity model adds recovery score† to the primary model predictors. Standardized coefficients ($\beta$) reported.

| Model | Predictor | $\beta$ | $p$ | $N$ | Adj. $R^2$ |
|---|---|---|---|---|---|
| Primary | BMI† | 0.687 | <0.001 | 379 | 0.301 |
| | Sleep duration | 0.063 | 0.657 | | |
| | Sleep efficiency | $-0.409$ | 0.005 | | |
| | Gender (female) | $-1.377$ | <0.001 | | |
| | Age | $-0.022$ | 0.877 | | |
| Sensitivity | BMI† | 0.497 | <0.001 | 379 | 0.359 |
| | Sleep duration | 0.181 | 0.186 | | |
| | Sleep efficiency | $-0.070$ | 0.638 | | |
| | Recovery score† | $-0.850$ | <0.001 | | |
| | Gender (female) | $-1.382$ | <0.001 | | |
| | Age | 0.109 | 0.430 | | |

† Indirect constituent; partly mediated through the algorithm.

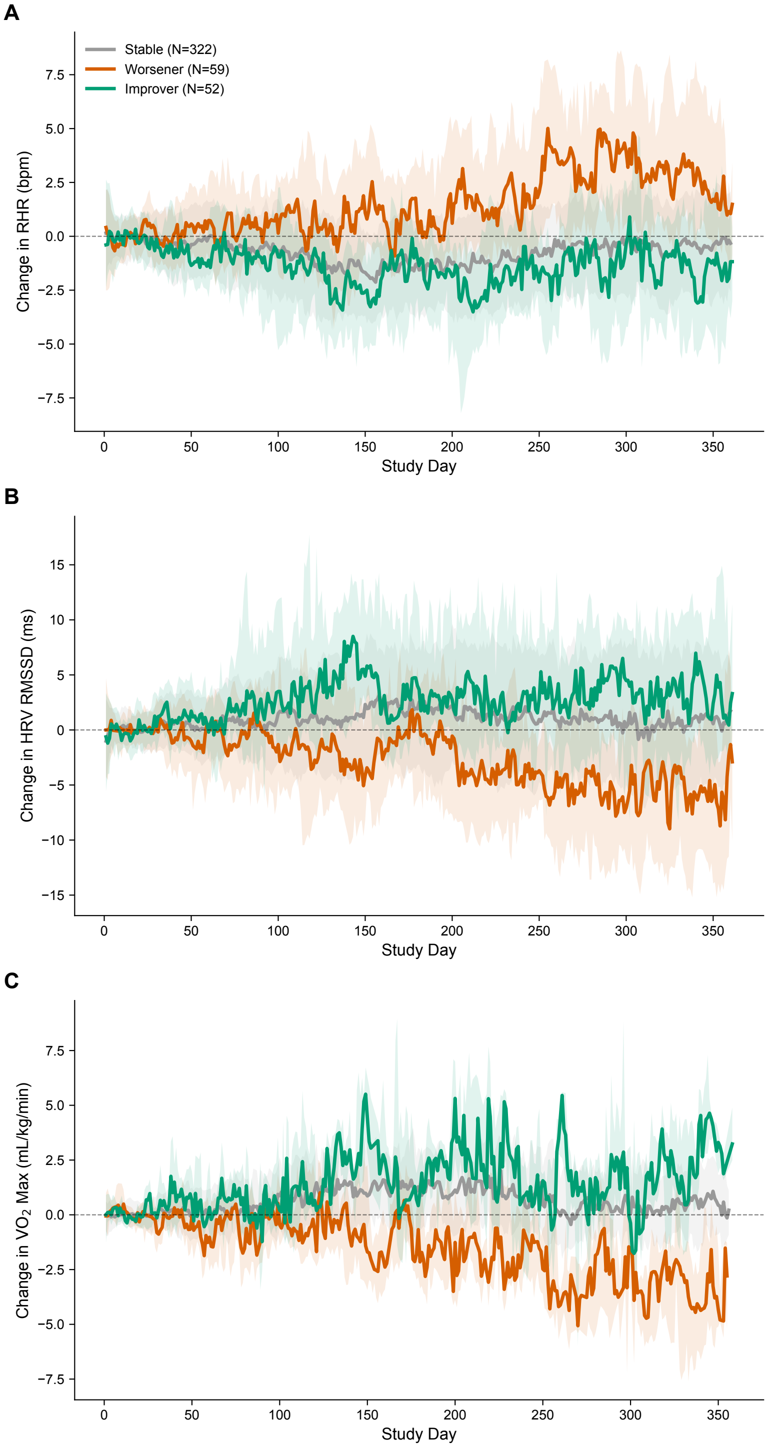


*All three metrics are constituent inputs to the Cardio Age algorithm. Daily median ± IQR (7-day rolling, baseline = first 30-day mean).*

**Figure S1.** Temporal divergence of constituent metrics between 12-month trajectory groups. Three panels show change from 30-day baseline in nighttime resting heart rate (top), HRV RMSSD (middle), and estimated $VO_2$ max (bottom) for sustained improvers ($N = 52$, green) and sustained worseners ($N = 59$, red). Divergence in resting heart rate and estimated $VO_2$ max becomes visible by months 3–4. All three metrics are direct constituent inputs to the Cardio Age algorithm; associations with Cardio Age trajectories are therefore partly tautological.
